# Physiological Perspective of Musculoskeletal Disorders among Older Peoples in Bangladesh

**DOI:** 10.1101/2023.07.17.23292700

**Authors:** Sharmila Jahan, Md. Rabiul Islam, Tania Rahman, Md. Feroz Kabir, Md. Kabir Hossain, K M Amran Hossain, Md. Zahid Hossain, Ehsanur Rahman, Md. Waliul Islam, Sonjit Kumar Chakrovorty, Altaf Hossain Sarker, Golam Moula, Atqiya Antara

**Author notes:** Corresponding Author Sharmila Jahan, Assistant Professor, Department of Physiotherapy and Rehabilitation, Faculty of Health Sciences, Jashore University of Science and Technology, Bangladesh.

## Abstract

Musculoskeletal disorders are debilitating conditions that significantly impact on the state of health, especially in elderly people. The study, which employed a cross-sectional design and practical sampling, included 206 participants from all over Bangladesh with musculoskeletal issues of varying severity and impact. The study was carried out between January and June of 2022. The majority of them experienced musculoskeletal pain. It was also common to have limited mobility as a result of arthritic change, which eventually affected daily activities like taking care of oneself. To improve the health of the elderly female population, more study must be conducted to identify the many factors that contribute to musculoskeletal issues. The development of effective prevention and rehabilitation programs must then be based on this knowledge.

## Background

Decreased adult mortality and fertility rates are responsible for the global ageing population [1]. The nineteenth-century demographic transition in affluent countries is currently reshaping society in developing and low-income countries [2]. Bangladesh has entered the third phase of demographic transition from a high mortality-high fertility regime to a low mortality-low fertility one [3]. The “demographic dividend,” where developing countries’ working-age population outnumbers their dependent age population, is an opportunity from this population transition. Diseases have shifted from communicable to non-communicable, especially chronic musculoskeletal ailments, as the population ages. Musculoskeletal problems were the most common chronic non-communicable disease in Brazil [4]. Chronic musculoskeletal problems affect quality of life, independence, and social involvement because pain is the main complaint [5]. Musculoskeletal discomfort costs second only to cardiovascular illness [6]. Low back pain, common in middle-aged and older people, affects work impairment, absenteeism, and costs [7]. “Nonspecific pain,” even when confined to a specific place (e.g., lower back), is common [8]. Healthcare providers struggle with diagnosis and treatment complexity. To create effective health policies that promote older health and prevent disabilities, one must understand the issue. This study examines musculoskeletal diseases in older Bangladeshis from a physiological standpoint. Musculoskeletal disorders and disabilities affect the elderly worldwide, justifying our investigation. Musculoskeletal problems are associated with mobility loss in elderly adults worldwide. Healthcare systems should prioritize reversible senior physical impairment. It’s easier to handle common musculoskeletal problems in Bangladesh’s elderly by understanding their prevalence and pattern. In Bangladesh, where aged care is scarce, family members may view older people as invalid and without access to vital interventions, causing them to suffer till death. Psychosocial factors can affect geriatric musculoskeletal ailments. Thus, understanding the physiological basis of prevalent musculoskeletal problems in the elderly is essential for optimizing management, lowering impairment, boosting independence, and improving quality of life. Objectives The goal of this study is to gain a better understanding of musculoskeletal problems from a physiological standpoint among Bangladeshi seniors.

## Methodology

With an emphasis on disability status and quality of life, as well as demographic, biological, psychological, and social factors, this study aimed to gain a physiological understanding of musculoskeletal problems in older persons (60 years) in Bangladesh. A total of 206 people from all around Bangladesh with musculoskeletal problems of various severity and impact participated in the study, which used a cross-sectional design and convenient sampling. Research was conducted between the months of January and June of 2022. Data was collected through interviews that covered topics such as participants’ demographics, the participants’ experiences with and viewpoints on musculoskeletal illnesses, the participants’ capacities to do daily tasks, and their overall quality of life. Information was gathered via in-person interviews and careful observation. Various statistical methods, including frequency and percentage distributions, as well as qualitative and quantitative analysis in tabular form, were applied to the data collection. Quality of life in the elderly was evaluated using the WHO Quality of Life Scale (WHOQOL-BREF). The IRB, the Bangladesh Physiotherapy Association (BPA), and participant informed permission all played roles in ensuring the study complied with ethical standards. All participants were free to leave the study at any moment without penalty. People over the age of 60 with musculoskeletal diseases who did not match the inclusion criteria or who could not provide informed permission were not considered for participation. Hospitals and rehabilitation clinics all around Bangladesh helped fund the study, which was run by the Institute of Social Welfare and Research at the University of Dhaka. In conclusion, this study contributes significantly to our understanding of musculoskeletal problems from a physiological standpoint among the elderly population in Bangladesh.

## Results

Table 1 provides socio-demographic characteristics of study participants. The table shows participants’ age, gender, dwelling area, marital status, Covid-19 status, and monthly income. Participants and variable percentages are shown. Most participants (60.7%) were 60–65 years old and married (88.3%).

**Table 1:** Socio-demographic characteristics of the participants.

| Variable | Total |  | Male |  | Female |  |
| --- | --- | --- | --- | --- | --- | --- |
|  | n | % | n | % | n | % |
| **Age (years)** |  |  |  |  |  |  |
| 60-65 | 125 | 60.7 | 71 | 57.3 | 54 | 65.9 |
| 66-70 | 64 | 31.1 | 42 | 33.9 | 22 | 26.8 |
| 71-75 | 13 | 6.3 | 10 | 8.1 | 3 | 3.7 |
| 76-80 | 4 | 1.9 | 1 | 0.8 | 3 | 3.7 |
| **Living Area** |  |  |  |  |  |  |
| Semi-Rural | 76 | 36.9 | 15 | 12.1 | 10 | 12.2 |
| Semi Urban | 25 | 12.1 | 18 | 14.5 | 22 | 26.8 |
| Rural | 40 | 19.4 | 44 | 35.5 | 32 | 39 |
| Urban | 65 | 31.6 | 47 | 37.9 | 18 | 22 |
| **Marital Status** |  |  |  |  |  |  |
| Single | 7 | 3.4 | 4 | 3.2 | 3 | 3.7 |
| Married | 182 | 88.3 | 115 | 92.7 | 67 | 81.7 |
| Married Single | 4 | 1.9 | 1 | 0.8 | 3 | 3.7 |
| Widow | 7 | 3.4 | 0 | 0 | 7 | 6.1 |
| Divorce | 3 | 1.5 | 0 | 0 | 3 | 3.7 |
| Separated | 2 | 1 | 2 | 1.6 | 0 | 0 |
| **Covid-Status** |  |  |  |  |  |  |
| Yes | 37 | 18 | 20 | 16.1 | 17 | 20.7 |
| No | 169 | 82 | 104 | 83.9 | 65 | 79.3 |
| **Monthly Income** |  |  |  |  |  |  |
| <10000 | 84 | 40.8 | 44 | 35.5 | 40 | 48.8 |
| 10000-24999 | 47 | 22.8 | 29 | 23.4 | 18 | 22 |
| 25000-49999 | 26 | 12.6 | 18 | 14.5 | 8 | 9.8 |
| 50000-99999 | 19 | 9.2 | 11 | 8.9 | 8 | 9.8 |
| ≥100000 | 30 | 14.6 | 22 | 17.7 | 8 | 9.8 |

The gender distribution of participants in the table can be used to determine if males and females differ significantly. Table 2 shows participant musculoskeletal problems. The table lists back, neck, knee, heel, hip, gross muscle, elbow, wrist, and fracture statistics. The number of participants who reported each ailment and gender breakdown are shown. The table also includes Chi-square values and p-values to investigate gender and musculoskeletal problems. Figure 1 shows the mean ADL score of individuals by comorbidity.

**Table 2:** Musculoskeletal Disorder among participants.

| **Condition** | **Total** |  | **Male** |  | **Female** |  | **X<sup>2</sup>** | **P-value** |
| --- | --- | --- | --- | --- | --- | --- | --- | --- |
|  | **n** | **%** | **n** | **%** | **n** | **%** |  |  |
| **Back pain** | 159 | 77.6% | 92 | 74.8% | 67 | 81.7% | 1.582 | 0.208 |
| **Neck pain** | 84 | 41.0% | 53 | 43.1% | 31 | 37.8% | 0.498 | 0.48 |
| **knee pain** | 173 | 84.4% | 100 | 81.3% | 73 | 89.0% | 2.576 | 0.108 |
| **Heel pain** | 23 | 11.2% | 14 | 11.4% | 9 | 11.0% | 0.005 | 0.944 |
| **Hip pain** | 13 | 6.3% | 8 | 6.5% | 5 | 6.1% | 0.01 | 0.919 |
| **Gross Muscle Pain** | 26 | 12.7% | 19 | 15.4% | 7 | 8.5% | 2.061 | 0.151 |
| **Elbow Pain** | 39 | 19.0% | 26 | 21.1% | 13 | 15.9% | 0.841 | 0.359 |
| **Wrist Pain** | 52 | 25.4% | 27 | 22.0% | 25 | 30.5% | 1.986 | 0.159 |
| **Fracture** | 4 | 2.0% | 4 | 3.3% | 0 | 0.0% |  |  |
| **Shoulder Pain** | 30 | 14.6% | 17 | 13.8% | 13 | 15.9% | 0.182 | 0.669 |

**Figure 1:**
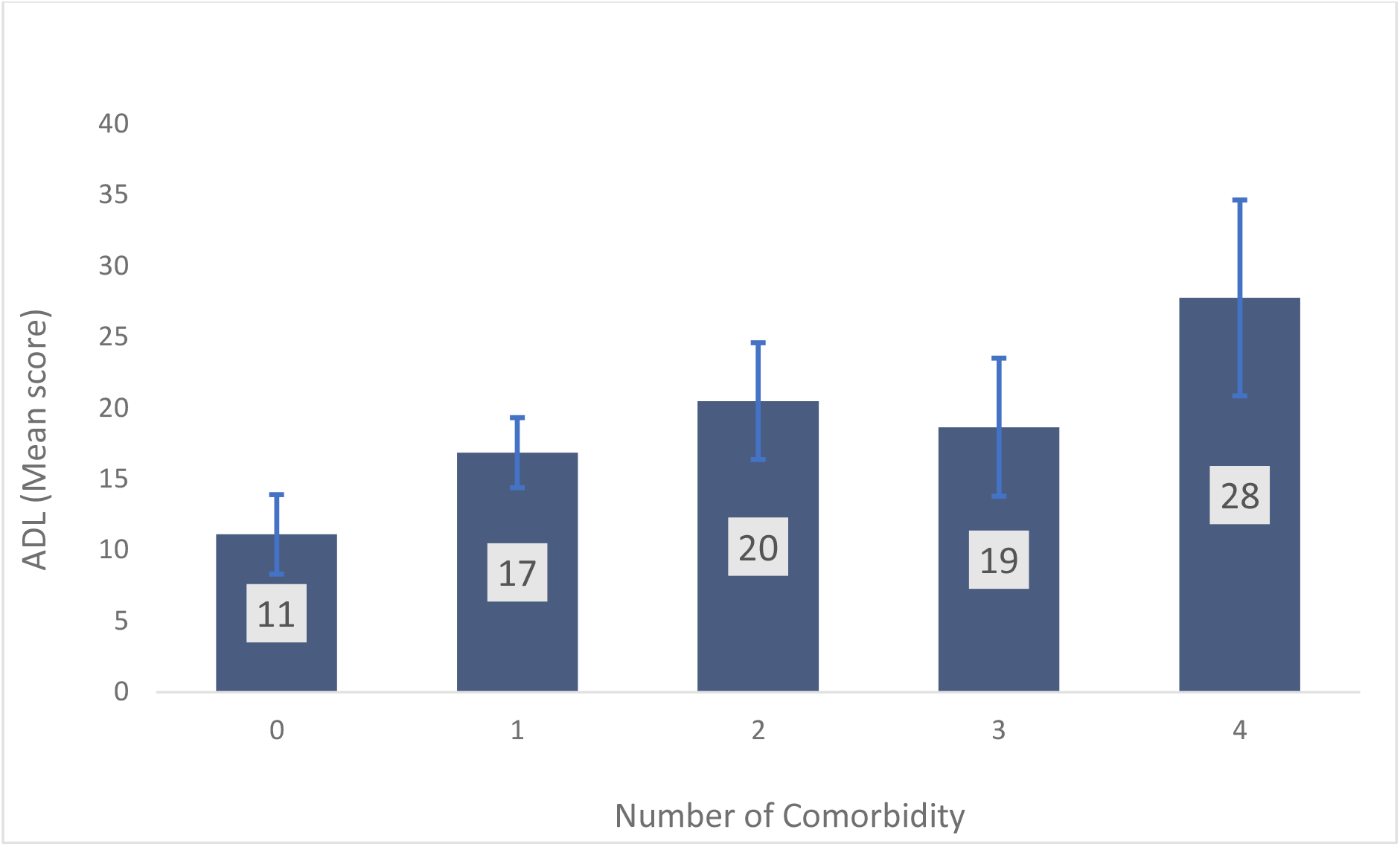
Mean ADL score of the Participants according to Number of Comorbidity

**Figure 1** for indicating that elderly person with 4 comorbidities has highest score for activity limitation. Were person with no comorbidity has only mean score of activity limitation 11.

The chart illustrates that people with four comorbidities had the highest activity limitation score, while those without any had a mean score of 11. The more comorbidities, the more activity constraint. Table 3 shows the association between quality of life and activities of daily living. The table shows Pearson’s correlation coefficients between WHO-QoL BREF scores for physical, psychological, social connection, and environmental domains and basic, instrumental, and typical social roles. Correlation coefficients measure two variables’ strength and direction. The table indicates a substantial positive correlation (0.95) between basic and instrumental activities of daily life, demonstrating that a person who can perform basic activities can also perform instrumental ones. The table also demonstrates negative associations between ADL measurements and physical and psychological domains of QoL scores, suggesting that increased activity limitation is related with lower QoL.

**Table 3:**
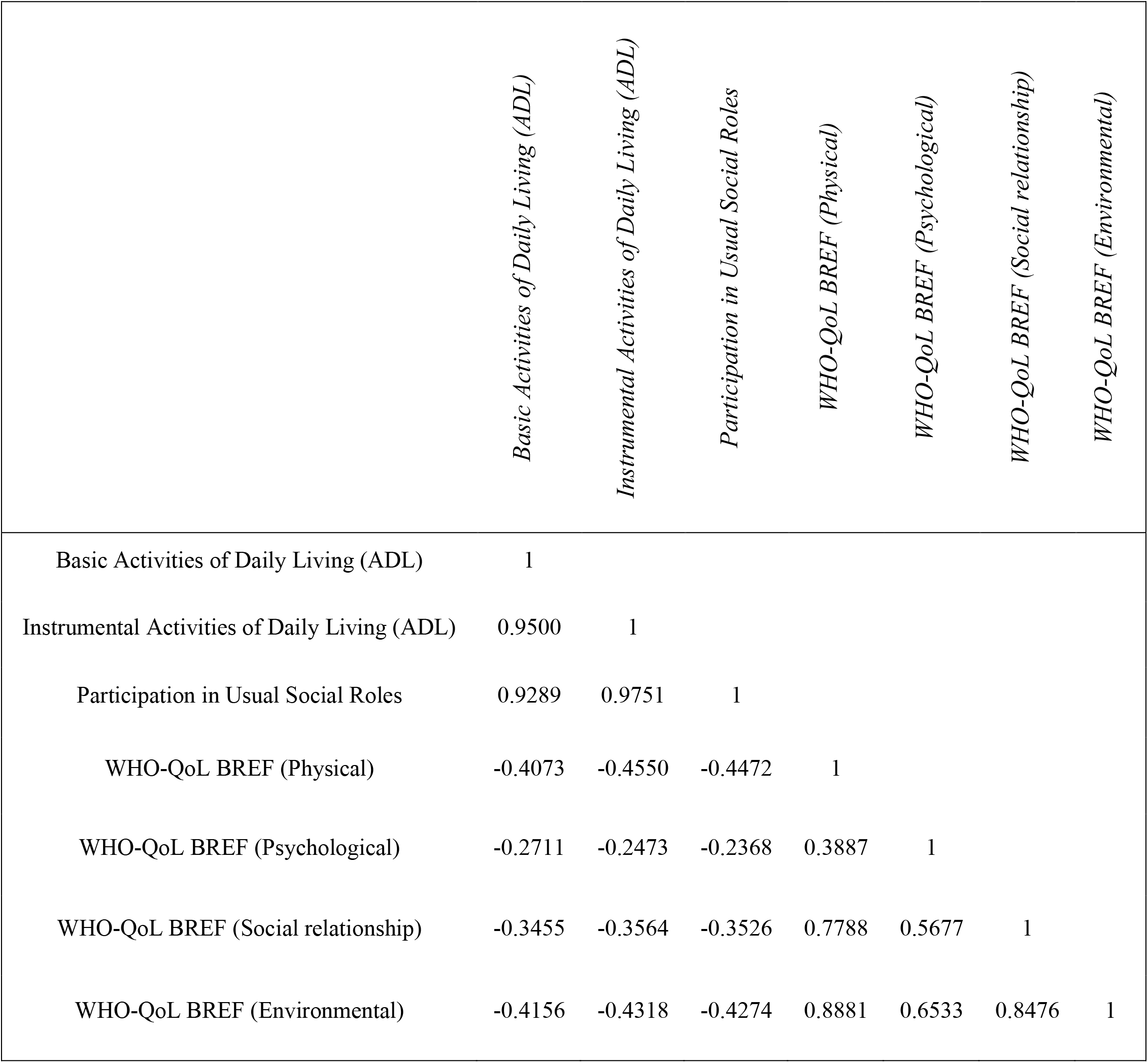
Correlation among Activity of daily living scale and Quality of life (Preasons Correlation)

## Discussion

Elderly people suffer from musculoskeletal problems that cause pain and disability. Neck, hip, knee, and lower back pain were reported by 98% of older persons [9]. One-third had knee joint restrictions. 84% of respondents had weak muscles and 78% struggled with daily [10]. Musculoskeletal and neurological problems that decrease muscle function caused geriatric falls. Physical disability, musculoskeletal issues, and inactivity increased falls. Western European and Japanese populations are aging, raising healthcare problems. Study heterogeneity affects older musculoskeletal disorder prevalence. Women report musculoskeletal pain more than males. Prevalence estimates tend to drop in the oldest age group (80+) [9]. Healthcare practitioners must learn more about older people’s health and disabilities due to their growing numbers. Most elderly people complained of joint pain, especially knee pain. Low back, shoulder, and neck pain were common. Over 70% of seniors report discomfort [11]. Southern India’s third most common complaint was back and neck pain. Elderly musculoskeletal issues included joint degeneration and restricted range of motion [12]. Hip, lower back, neck, shoulder, and ankle joints were the most restricted. Knee pain was the most common complaint, followed by low back, neck, shoulder, and ankle discomfort. Joint mobility and pain were linked [13]. Older folks, especially ladies, were disabled by arthritis. Knee pain was the most common musculoskeletal symptom in elderly women in this study, at 84%. 25% of subjects reported wrist pain. Shoulder pain was reported by 14.6% of participants, but neck pain by 50% [14]. Optimal healthcare is needed to keep elderly people functional. Musculoskeletal problems in the aging population must be better understood due to their rising prevalence.

## Conclusion

Nearly all of them had musculoskeletal pain. It was also typical to have restricted mobility due to arthritic change, which eventually has an impact on daily tasks like self-care. More research must be done to uncover the numerous elements that link to musculoskeletal problems in order to enhance the health of the female aged population. This research must then be used to create successful programs for both prevention and rehabilitation.

## Data Availability

All data produced in the present study are available upon reasonable request to the authors

## Ethical consideration

The proposal was approved by the Institutional Review Board (IRB) of the Institute of Physiotherapy, Rehabilitation, and Research of Bangladesh Physiotherapy Association (BPA-IPRR/IRB/19/01/2021/65). Written consent was taken from each participant before collecting the data. Informed consent was taken verbally during the phone call and written during the household survey. To ensure confidentiality, ethics, and privacy, the Declaration of Helsinki principles (15) were maintained throughout the research. One of our research team members obtained household screening approval from the Directorate General of Health Services of the Government of the People’s Republic of Bangladesh.

